# The interaction between social determinants of health, health behaviors, and child’s intellectual developmental diagnosis

**DOI:** 10.1101/2024.05.23.24307804

**Authors:** Phoebe P. Tchoua, Emily Clarke, Heather Wasser, Seema Agrawal, Rebecca Scothorn, Kelsey Thompson, Michaela Schenkelberg, Erik A. Willis

## Abstract

**INTRODUCTION:** Social determinants of health (SDOH) may impact caregivers’ ability to implement evidence-based health practices at home during early childhood, especially in families with children with intellectual and developmental disabilities (IDD). Therefore, we examined the influence of SDOH and children’s diagnosis (typically developing [TD], Down syndrome [DS], autism) on caregiver’s self-report of meeting evidence-based health practices.

**METHODS:** Caregivers (n=172) of children ages 2-6 years (TD: n=93, DS: n=40, autism: n=39) completed an online survey on SDOH and health practices related to child nutrition (CN), physical activity (PA), outdoor play (OP), and screen time (ST). A total SDOH score was computed by assigning 1 point for each favorable SDOH metric (range 0-13). Linear regressions were used to examine associations between SDOH and CN, PA, OP, ST health practices and the moderating effect of IDD diagnosis.

**RESULTS:** Most caregivers were non-Hispanic White (84.3%), female (76.7%), 18-35 years old (55.2%), and married (89.5%). The DS group had the lowest SDOH score (mean = 8.4±1.0) compared to autism (mean = 10.1±1.0) and TD (mean = 11.0±0.9). No family scored 100% in evidence-based practices for any health practice. SDOH score was significantly associated with evidence-based practices met score for CN (b = 1.94, 95% CI = 0.84, 3.04; p = 0.001) and PA (b = 4.86, 95% CI = 2.92, 6.79; p <0.0001). Moderation analysis showed no association in the DS and autism groups between SDOH score and CN percent total score, or between SDOH score and CN, PA, and OP for percent evidence-based practices met. SDOH score was also not associated with OP percent total score for the DS group.

**CONCLUSIONS:** This study highlights the differential influence of SDOH on caregivers’ implementing health practices in families with children of different IDD diagnoses. Future research is needed to understand impacts of SDOH on non-typically developing children.

## Introduction

> Note: Person-first language is not the preferred choice for all communities. To avoid confusion for the reader, person-first language will be used to refer to children with Down syndrome as ‘children with Down syndrome,’ and identity-first language for children with an autism spectrum disorder diagnosis as ‘autistic children’ throughout the manuscript.

There is a growing recognition that parents, caregivers, parenting practices, and the home environment play a critical role in shaping lifelong health behaviors such as healthy eating, physical activity (PA), and screen time during early childhood. Research shows a significant association between food parenting practices, parental education level, and children’s dietary intake. Children with parents of high education level are more likely to eat more fruits and vegetables and be physically active compared with children of parents of low education level (1, 2). Parents are also instrumental in helping children be more active through modeling active behavior and creating a home environment that promotes PA and reduces screen time (3). Ultimately, the influence of parents and caregivers during early childhood lays the foundation for lifelong health behaviors. Yet, young children struggle to meet evidence-based recommendations regarding nutrition, PA, and screen time (4–9).

However, while parents and caregivers have a significant immediate and long-term impact on their child’s wellbeing, it is important to recognize that social determinants of health (SDOH) such as socioeconomics, environmental context, community factors and individual children’s developmental abilities significantly influence caregiver’s capacity to consistently promote good health behaviors. Lower caregiver educational level, higher rates of poverty, and neighborhoods with unfavorable conditions (e.g., safety concerns, limited or no sidewalks/walking paths) have been associated with higher rates of obesity and physical inactivity in children (10, 11). Specifically, poverty exposure prior to age two is strongly associated with childhood obesity (12) and negatively associated with children’s overall health (13). Additionally, children of parents with less than a high school education are more likely to reside in unsafe neighborhoods that lack essential health-promoting features like sidewalks, parks, playgrounds, and recreation centers, and have 30-60 percent higher likelihood of childhood obesity (10). Furthermore, SDOH contribute to more health inequities and challenges in individuals with intellectual and developmental disabilities (IDD; e.g., Down syndrome, autism) (14), who historically have been underrepresented in health behavior research, widening the health disparity gap between children with and without IDD (15). Children with IDD experience difficulties in activities of daily living, motor skills, communication, and participation at home, school, and in the community (10, 11), and are more susceptible to physical inactivity and obesity (10, 11, 16–18). Caregivers of children with IDD face distinct challenges, including the need for more intensive interventions, coordination among multiple healthcare providers, management of co-morbidities, and navigating the absence of standardized treatments (14). These families often encounter significant financial burdens due to healthcare costs, medication costs, not being able to work, supplemental therapies and other support services which highlights the multifaceted demands of caregiving in this population (14, 19). Understanding how these additional challenges impact parents’ and caregivers’ ability to promote health behaviors are essential steps toward fostering healthier environments and improving outcomes for all children, especially those with IDD.

Existing studies have explored the impact of SDOH on health and parenting styles (20–22), however, to our knowledge, no study has investigated the relationships between SDOH and caregivers’ evidence-based health practices within the home environment, particularly among families with children with IDD. In the current study we leveraged data obtained from a web-based family health practices survey to investigate the influence of SDOH on caregiver practices related to child nutrition, PA, outdoor play, and screen time in families with typically developing (TD) children, those with Down syndrome (DS), and autistic children, aged 2-6 years. This study seeks to gain insights into the underlying external factors (i.e., SDOH factors) influencing health behaviors within diverse family contexts, informing strategies for promoting healthier lifestyles among all children.

## Methods

### Study design and participants

Our study sample comprised participants who completed a web-based cross-sectional survey to explore family-level health practices around seven content areas: 1) physical activity, 2) outdoor play and learning, 3) child nutrition (2-6 years only), 4) oral health, 5) farm to home (i.e., serve family local foods for meals or snacks, gardening and gardening activities with child, education about fresh and local foods), 6) screen time, and 7) breast/infant feeding (0-2 years only). The survey also included questions about the types of health information resources used and participants’ trust in these resources. The researchers employed convenience sampling to recruit parents and caregivers of children aged 0-6 years old with DS, autism, or TD to complete the online survey. Participants were recruited between November 2022 and February 2023 through flyers, advertisements in traditional social media platforms (LinkedIn, X [formerly Twitter], Facebook, Reddit) and emails to relevant foundations and organizations. To be eligible for the survey, respondents were required to be 18 years or older, live in the United States, and have a child aged 0-6 years. Participants were excluded if their child required a full liquid, mechanically altered, soft, pureed, and/or tube fed diet. Participants provided consent before taking the survey and were eligible to receive one of thirty randomly allocated, $50 gift cards after completion. More details on the survey development and participant recruitment can be found elsewhere Thompson, Clarke (23). This manuscript was written in alignment with the checklist for the Strengthening the Reporting of Observational Studies in Epidemiology (STROBE) Statement. The study was reviewed by the University of North Carolina at Chapel Hill Institutional Review Board and was determined exempt.

The present study focused on four family-level health practice content areas for children ages 2 to 6 years: child nutrition, PA, outdoor play and learning (hereinafter, outdoor play), and screen time practices. Of the 659 primary caregivers who were eligible and consented to participate, 387 completed the survey. We excluded participants whose child was aged 2 years or younger (n = 171), and those who were missing relevant data necessary for calculating SDOH metric scores (n = 44).

### Measures

#### Demographics

Caregivers self-reported basic demographic information (age, race/ethnicity, sex, marital status, education level, household income) in the online survey. Education level was reported as “Less than high school”, “High school graduate or GED”, “Some college or technical school”, “Associate degree”, “Bachelor degree”, or “Graduate or professional degree” and household income was reported as “less than $15,000”; “$15,000 to $34,999”; “$35,000 to $74,999”; “$75,000 to $149,999”; or “$150,000 or more.” Caregivers also self-reported their child’s age group (0 −12 months, 12-23 months, 2 to 6 years) and developmental disability diagnosis (TD, DS, autism).

#### SDOH

All SDOH metrics and their corresponding survey items are described in Table 1. Caregivers self-reported on 13 items related to SDOH, which were grouped into five broad SDOH metrics, in line with the definitions provided by the Office of Disease Prevention and Health Promotion’s Healthy People 2030 and the World Health Organization: Economic Stability, Education Access and Quality, Healthcare Access, Residential Environments, and Social Context and Support (22).

**Table 1.**
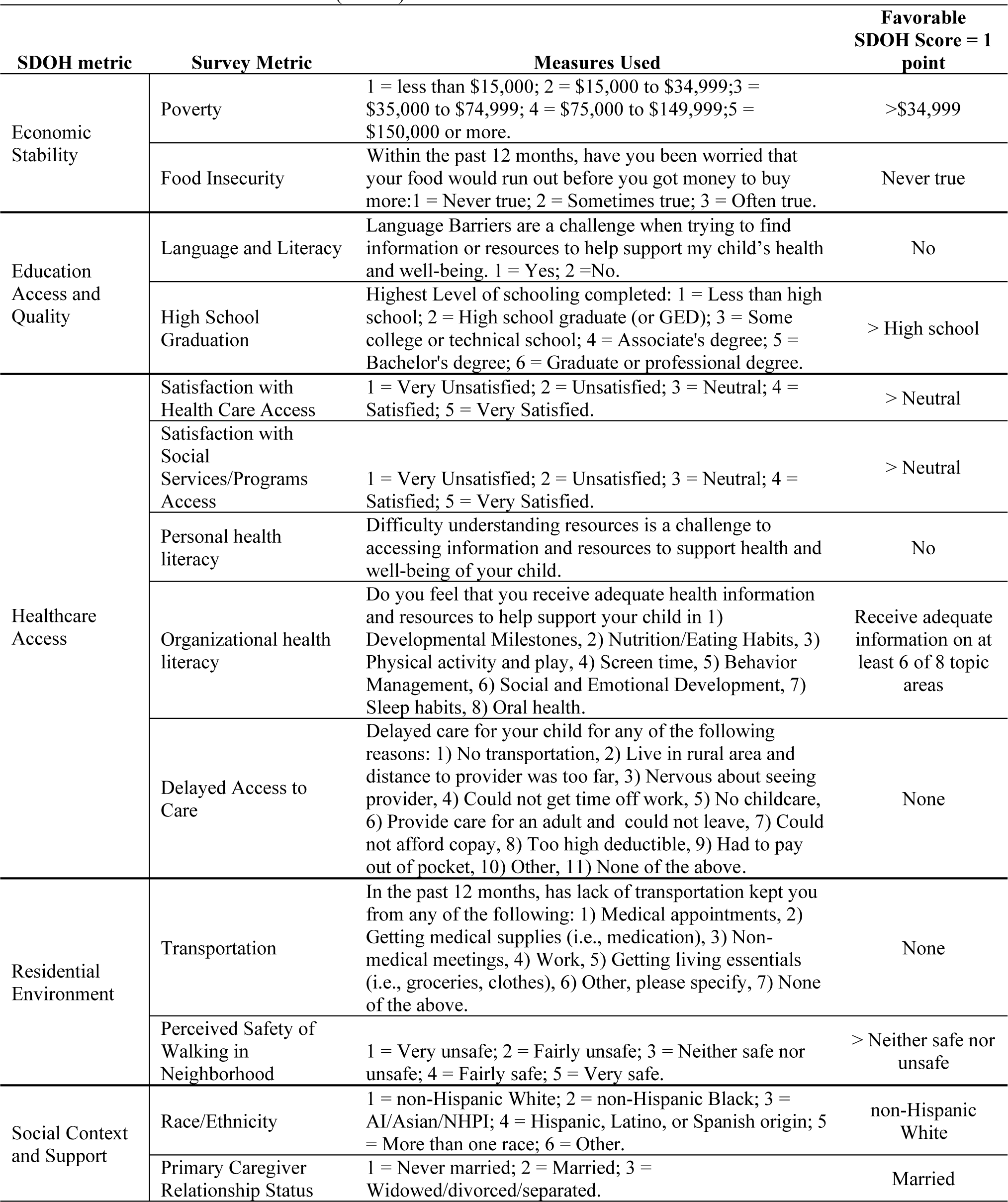
Social determinants of health (SDOH) metric definitions.

| SDOH metric | Survey Metric | Measures Used | Favorable SDOH Score = 1 point |
| --- | --- | --- | --- |
| Economic Stability | Poverty | 1 = less than \$15,000; 2 = \$15,000 to \$34,999; 3 = \$35,000 to \$74,999; 4 = \$75,000 to \$149,999; 5 = \$150,000 or more. | >\$34,999 |
|  | Food Insecurity | Within the past 12 months, have you been worried that your food would run out before you got money to buy more: 1 = Never true; 2 = Sometimes true; 3 = Often true. | Never true |
| Education Access and Quality | Language and Literacy | Language Barriers are a challenge when trying to find information or resources to help support my child's health and well-being. 1 = Yes; 2 = No. | No |
|  | High School Graduation | Highest Level of schooling completed: 1 = Less than high school; 2 = High school graduate (or GED); 3 = Some college or technical school; 4 = Associate's degree; 5 = Bachelor's degree; 6 = Graduate or professional degree. | > High school |
| Healthcare Access | Satisfaction with Health Care Access | 1 = Very Unsatisfied; 2 = Unsatisfied; 3 = Neutral; 4 = Satisfied; 5 = Very Satisfied. | > Neutral |
|  | Satisfaction with Social Services/Programs Access | 1 = Very Unsatisfied; 2 = Unsatisfied; 3 = Neutral; 4 = Satisfied; 5 = Very Satisfied. | > Neutral |
|  | Personal health literacy | Difficulty understanding resources is a challenge to accessing information and resources to support health and well-being of your child. | No |
|  | Organizational health literacy | Do you feel that you receive adequate health information and resources to help support your child in 1) Developmental Milestones, 2) Nutrition/Eating Habits, 3) Physical activity and play, 4) Screen time, 5) Behavior Management, 6) Social and Emotional Development, 7) Sleep habits, 8) Oral health. | Receive adequate information on at least 6 of 8 topic areas |
|  | Delayed Access to Care | Delayed care for your child for any of the following reasons: 1) No transportation, 2) Live in rural area and distance to provider was too far, 3) Nervous about seeing provider, 4) Could not get time off work, 5) No childcare, 6) Provide care for an adult and could not leave, 7) Could not afford copay, 8) Too high deductible, 9) Had to pay out of pocket, 10) Other, 11) None of the above. | None |
| Residential Environment | Transportation | In the past 12 months, has lack of transportation kept you from any of the following: 1) Medical appointments, 2) Getting medical supplies (i.e., medication), 3) Non-medical meetings, 4) Work, 5) Getting living essentials (i.e., groceries, clothes), 6) Other, please specify, 7) None of the above. | None |
|  | Perceived Safety of Walking in Neighborhood | 1 = Very unsafe; 2 = Fairly unsafe; 3 = Neither safe nor unsafe; 4 = Fairly safe; 5 = Very safe. | > Neither safe nor unsafe |
| Social Context and Support | Race/Ethnicity | 1 = non-Hispanic White; 2 = non-Hispanic Black; 3 = AI/Asian/NHPI; 4 = Hispanic, Latino, or Spanish origin; 5 = More than one race; 6 = Other. | non-Hispanic White |
|  | Primary Caregiver Relationship Status | 1 = Never married; 2 = Married; 3 = Widowed/divorced/separated. | Married |

The Economic Stability score was computed based on total household income (favorable score >$34,999) and the perception of food insecurity (favorable score = never true), resulting in a construct score ranging from 0 (no favorable SDOH) to 2 (all favorable SDOH) points.

Education Access and Quality score was determined based on language and literacy barriers (favorable score = no) and education level (favorable score = greater than a high school education), with a construct score range from 0 to 2 points.

The Healthcare Access score was derived from satisfaction with healthcare access (favorable score = at least satisfied), satisfaction with social services/programs access (favorable score = at least satisfied), personal health literacy (favorable score = no difficulty), organizational health literacy (favorable score = receive adequate information on at least 6 of 8 wellness-related topic areas), and barriers to care (favorable score = no barriers reported), with a construct score range from 0 to 5 points.

Residential Environment score was calculated based on transportation issues (favorable score = none) and perceived neighborhood safety (favorable score = at least fairly safe), resulting in a construct score range from 0 to 2 points. Social Context and Support score was determined based on race and ethnicity (favorable score = non-Hispanic white) and relationship status (favorable score = married living together), with a construct score range from 0 to 2 points.

A total SDOH score was computed by assigning 1 point for each favorable survey metric, resulting in a range from 0 (no favorable SDOH) to 13 (all favorable SDOH). Each variable was weighted equally, consistent with prior analyses (24–26).

#### Family-level health practices

Self-assessments were developed for each of the four content areas using questionnaire items adapted for home use from the web-based Nutrition and Physical Activity Self-Assessment for Child Care’s (Go NAPSACC) family child care home assessment tool (27). The Go NAPSACC family child care home self-assessment items allow individuals who provide care for children in their homes to evaluate to what degree they are meeting current evidence-based recommendations. These self-assessments were created following a thorough review of regulations, performance standards, and recommendations from scientific literature and governmental and professional organizations. Survey adaptation was a collaborative effort among experts in early childhood health behaviors, registered dietitians, and key informants. Final items were reviewed by experts for face validity prior to dissemination. Final self-assessments included 35 items for child nutrition, 14 items for PA, 10 items for outdoor play, and 6 items for screen use.

Survey items were scored using a 4-point Likert-type scale, from 1 = “not engaging”, 2 = “minimally engaging”, 3 = “somewhat engaging”, to 4 = “fully engaging” in evidence-based practice recommendations. Self-assessment total percentage scores (total percentage score) were calculated for each content area by summing all scored items divided by the total possible points for all applicable items multiplied by 100, yielding a percentage score between 0 (least engaging) to 100 (most engaging). The percent of evidence-based practices met for each topic area was calculated by summing the number of items where the evidence-based practice was fully engaged (score = 4) and dividing this number by the total number of evidence-based practices multiplied by 100, yielding a percentage score between 0 (not fully engaging) to 100 (fully engaging). The total percentage and evidence-based practice met percentage score helps measure the full range of caregivers’ engagement in evidence-based practices.

### Statistical analyses

Data were summarized using means and standard deviations for continuous variables, and frequencies and percentages for categorical variables. Separate linear regression models were used to examine direct associations between SDOH and evidence-based family-level health practice content areas (i.e., child nutrition, PA, outdoor play, screen time), as well as the moderating effect of a child diagnosis (TD, DS, autism). All models were adjusted for primary caregivers’ age, sex, and the child’s diagnosis. Interaction terms between child diagnosis and SDOH total score were entered into the models to examine potential moderating effects. Statistical significance was set at p = 0.05 for interpreting main effects and at p = 0.10 for interpreting moderating effects. All analyses were conducted using SAS version 9.4 (Cary, NC).

## Results

### Participants

The median completion time of the online survey for participants was 29 minutes. Table 2 shows the participant characteristics of the 172 primary caregivers that were included in this analysis. Overall, caregivers were primarily non-Hispanic White (84.3%), female (76.7%), 18-35 years old (55.2%), married (89.5%), held a graduate or professional degree (36.6%), and had a household income of $75,000 - $149,999 (47.1%). Fifty-four percent of families self-reported their child was TD, 23% reported their child was diagnosed with DS, and 23% reported their child was diagnosed with autism. Primary caregivers of a child with DS included in this study were younger (85.0% between 18-35 years old) compared to caregivers of TD and autistic children with 52.7% and 30.8% being between 18-35 years old, respectively. Additionally, male caregivers (47.5%) more frequently completed the survey for the children with DS, compared to the TD and (17.2%) and autism (12.8%) groups. Among autistic children, 74.4% of primary caregivers were married compared to 97.5% and 92.5% among caregivers of a child with DS and TD child, respectively.

**Table 2.** Sample characteristics and descriptive information of social determinants of health by child diagnosis.

| Variable | All parents<br>(n= 172) |  | Typically Developing<br>(n= 93) |  | Down syndrome<br>(n= 40) |  | Autism<br>(n= 39) |  |
| --- | --- | --- | --- | --- | --- | --- | --- | --- |
|  | n | % | n | % | n | % | n | % |
| **CAREGIVER CHARACTERISTICS** |  |  |  |  |  |  |  |  |
| **Age (years)** |  |  |  |  |  |  |  |  |
| *18 to 35* | 95 | 55.2 | 49 | 52.7 | 34 | 85 | 12 | 30.8 |
| *35 to 50* | 77 | 44.8 | 44 | 47.3 | 6 | 15 | 27 | 69.2 |
| **Biological sex** |  |  |  |  |  |  |  |  |
| *Female* | 132 | 76.7 | 77 | 82.8 | 21 | 52.5 | 34 | 87.2 |
| *Male* | 40 | 23.3 | 16 | 17.2 | 19 | 47.5 | 5 | 12.8 |
| **SDOH CHARACTERISTICS** |  |  |  |  |  |  |  |  |
| **Economic Stability** |  |  |  |  |  |  |  |  |
| **Total household income** |  |  |  |  |  |  |  |  |
| *≤\$34,999* | 16 | 9.3 | 7 | 7.5 | 3 | 7.5 | 6 | 15.4 |
| *≥\$35,000* | 156 | 90.7 | 86 | 92.5 | 37 | 92.5 | 33 | 84.6 |
| **Household food security level** |  |  |  |  |  |  |  |  |
| *Sometimes or often true* | 57 | 33.1 | 23 | 24.7 | 27 | 67.5 | 7 | 17.9 |
| *Never true* | 115 | 66.9 | 70 | 75.3 | 13 | 32.5 | 32 | 82.1 |
| **Education Access and Quality** |  |  |  |  |  |  |  |  |
| **Language barriers (no)** |  |  |  |  |  |  |  |  |
| *Yes* | 21 | 12.2 | 10 | 10.7 | 10 | 25 | 1 | 2.6 |
| *No* | 151 | 87.8 | 83 | 89.3 | 30 | 75 | 38 | 97.4 |
| **Primary caregiver education** |  |  |  |  |  |  |  |  |
| *≤ High School Graduate* | 7 | 4.1 | 3 | 3.2 | 1 | 2.5 | 3 | 7.7 |
| *>High School Graduate* | 165 | 95.9 | 90 | 96.8 | 39 | 97.5 | 36 | 92.3 |
| **Healthcare Access** |  |  |  |  |  |  |  |  |
| **Satisfaction with health care access** |  |  |  |  |  |  |  |  |
| *Unsatisfied/Neutral* | 40 | 23.4 | 14 | 15.1 | 10 | 25 | 16 | 41 |
| *Satisfied* | 132 | 76.7 | 79 | 84.9 | 30 | 75 | 23 | 59 |
| **Satisfaction with social services/programs access** |  |  |  |  |  |  |  |  |
| *Unsatisfied/Neutral* | 60 | 34.9 | 27 | 29 | 11 | 27.5 | 22 | 56.4 |
| *Satisfied* | 112 | 65.1 | 66 | 71 | 29 | 72.5 | 17 | 43.6 |
| **Difficulty understanding resources** |  |  |  |  |  |  |  |  |
| *Yes* | 34 | 19.8 | 10 | 10.7 | 22 | 55 | 2 | 5.1 |
| *No* | 138 | 80.2 | 83 | 89.3 | 18 | 45 | 37 | 94.9 |
| **Receive adequate health information and resources (Yes)** |  |  |  |  |  |  |  |  |
| *Developmental Milestones* | 137 | 79.7 | 80 | 86 | 27 | 67.5 | 30 | 76.9 |
| *Nutrition/Eating Habits* | 129 | 75 | 77 | 82.8 | 27 | 67.5 | 25 | 64.1 |
| *Physical activity and play* | 133 | 77.3 | 77 | 82.8 | 29 | 72.5 | 27 | 69.2 |
| *Screen time* | 135 | 78.5 | 78 | 83.9 | 26 | 65 | 31 | 79.5 |
| *Behavior Management* | 121 | 70.4 | 67 | 72 | 33 | 82.5 | 21 | 53.9 |
| *Social and Emotional Development* | 120 | 69.8 | 72 | 77.4 | 28 | 70 | 20 | 51.3 |
| *Sleep habits* | 132 | 76.7 | 76 | 81.7 | 33 | 82.5 | 23 | 59 |
| *Oral health* | 143 | 83.1 | 81 | 87.1 | 28 | 70 | 34 | 87.2 |

**Table 2.** (Continued) Sample characteristics and descriptive information of social determinants of health by child diagnosis.
| Variable | All parents<br>(n= 172) |  | Typically Developing<br>(n= 93) |  | Down syndrome<br>(n= 40) |  | Autism<br>(n= 39) |  |
| --- | --- | --- | --- | --- | --- | --- | --- | --- |
|  | n | % | n | % | n | % | n | % |
| **SDOH CHARACTERISTICS** |  |  |  |  |  |  |  |  |
| **Healthcare Access (Continued)** |  |  |  |  |  |  |  |  |
| **Have you delayed care for your child because (No)** |  |  |  |  |  |  |  |  |
| Did not have transportation | 160 | 93 | 89 | 95.7 | 33 | 82.5 | 38 | 97.4 |
| Distance to health care provider is too far | 153 | 89 | 88 | 94.6 | 28 | 70 | 37 | 94.9 |
| Nervous about seeing a health care provider | 146 | 84.9 | 83 | 89.3 | 25 | 62.5 | 38 | 97.4 |
| Could not get time off work | 154 | 89.5 | 89 | 95.7 | 31 | 77.5 | 34 | 87.2 |
| Could not get childcare | 155 | 90.1 | 85 | 91.4 | 33 | 82.5 | 37 | 94.9 |
| Provide care to an adult and could not leave them | 157 | 91.3 | 89 | 95.7 | 29 | 72.5 | 39 | 100 |
| Could not afford copay | 167 | 97.1 | 90 | 96.8 | 38 | 95 | 39 | 100 |
| Too high/or could not afford the deductible | 161 | 93.6 | 89 | 95.7 | 35 | 87.5 | 37 | 94.9 |
| Had to pay out of pocket for some/all of the procedure | 152 | 88.4 | 86 | 92.5 | 30 | 75 | 36 | 92.3 |
| Other, please specify | 167 | 97.1 | 93 | 100 | 40 | 100 | 34 | 87.2 |
| **Residential Environment** |  |  |  |  |  |  |  |  |
| **Lack of transportation kept you from (No)** |  |  |  |  |  |  |  |  |
| Medical appointments | 152 | 88.4 | 89 | 95.7 | 25 | 62.5 | 38 | 97.4 |
| Getting medical supplies (i.e., medication) | 151 | 87.8 | 84 | 90.3 | 29 | 72.5 | 38 | 97.4 |
| Non-medical meetings | 152 | 88.4 | 80 | 86 | 33 | 82.5 | 39 | 100 |
| Work | 148 | 86.1 | 85 | 91.4 | 25 | 62.5 | 38 | 97.4 |
| Getting living essentials (i.e., groceries, clothes) | 153 | 89 | 87 | 93.6 | 28 | 70 | 38 | 97.4 |
| Other, please specify | 172 | 100 | 93 | 100 | 40 | 100 | 39 | 100 |
| **Perceived safety of walking in neighborhood** |  |  |  |  |  |  |  |  |
| Unsafe/Neutral | 23 | 13.4 | 9 | 9.7 | 9 | 25.5 | 5 | 12.8 |
| Safe | 149 | 86.6 | 84 | 90.3 | 31 | 77.5 | 34 | 87.2 |
| **Social context and support** |  |  |  |  |  |  |  |  |
| **Race/Ethnicity** |  |  |  |  |  |  |  |  |
| non-white | 27 | 15.7 | 11 | 11.8 | 7 | 17.5 | 9 | 23.1 |
| non-Hispanic white | 145 | 84.3 | 82 | 88.2 | 33 | 82.5 | 30 | 76.9 |
| **Primary caregiver relationship status** |  |  |  |  |  |  |  |  |
| Never/Widowed/divorced/separated | 18 | 10.5 | 7 | 7.5 | 1 | 2.5 | 10 | 25.6 |
| Married | 154 | 89.5 | 86 | 92.5 | 39 | 97.5 | 29 | 74.4 |

### SDOH

The overall SDOH mean score was 10.2 points (standard deviation [SD] = 1.4). Total SDOH score was significantly lower for primary caregivers of a child with Down syndrome (8.4 points, SD = 1.0) compared to autistic children (10.1 points, SD = 1.0; p < 0.001) and those that are TD (11.0 points, SD = 0.9, p = 0.006). Primary caregivers of children with DS self-reported the highest prevalence of food insecurity (DS = 67.5% vs. TD = 24.7% vs. autism = 17.9%), language barriers related to health information and resources (DS = 25.0% vs. TD = 10.7% vs. autism = 2.6%), difficulty understanding resources (DS = 55.0% vs. TD = 10.7% vs. autism = 5.1%), transportation issues (DS = 85.0% vs. TD = 21.5% vs. autism = 5.1%), and delaying care for their child (DS = 87.5% vs. autism = 33.3% vs. TD = 23.6%; Table 2). Primary caregivers of autistic children reported the highest prevalence of dissatisfaction with access to healthcare services (autism = 41.0% vs. DS = 25.0% vs. TD = 15.1%), social services programs (autism = 56.4% vs. TD = 29.0% vs. DS = 27.5%), and health-related resources (autism = 48.7% vs. DS = 35.0% vs. TD = 24.7%; Table 2).

#### Association between SDOH and family-level health practices

Figure 1 shows the association between child nutrition (Figure 1A), PA (Figure 1B), outdoor play (Figure 1C), screen time (Figure 1D) evidence-based practices and SDOH metric score. Child nutrition total percentage score ranged 56.7% to 76.4%, with percent of evidence-based practice met percentage scores ranging from 0% to 45.6%. SDOH score was significantly associated with both the total percentage score (b = 0.81, 95% CI = 0.28, 1.33; p = 0.003) and the percent of evidence-based practices met (b = 1.94, 95% CI = 0.84, 3.04, p = 0.001) with child nutrition family practices.

**Figure 1.**
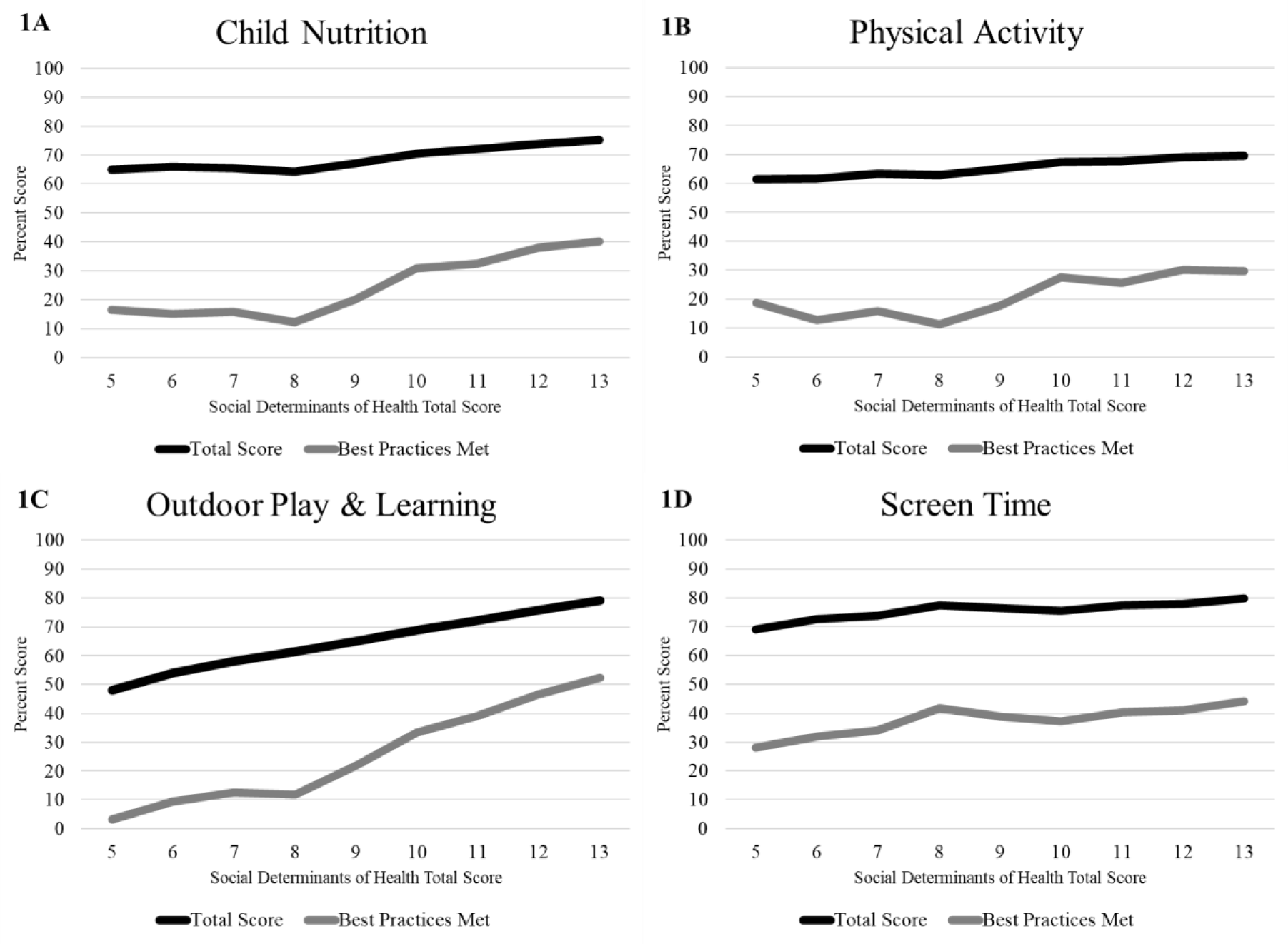
Association between child nutrition (Figure 1A), physical activity (Figure 1B), outdoor play and learning (Figure 1C), screen time (Figure 1D), self-assessment total percentage score and evidence-based practice met percentage score and SDOH metrics score. Total Score is calculated by summing all scored items divided by the total possible points for all applicable items multiplied by 100, yielding a percentage score between 0 (least engaging) to 100 (most engaging). Evidence-based practice met percentage score is calculated by summing the number of items where the best practice was fully engaged (score = 4) and dividing this number by the total number of evidence-based practice multiplied by 100, yielding a percentage score between 0 (not fully engaging) to 100 (fully engaging).

Similarly, PA total percentage scores varied between 55.9% and 73.6%, with evidence-based practice met percentage scores ranging from 0% to 40.3%. Here, the SDOH score was significantly associated with total percentage score (b = 1.10, 95% CI = 0.25, 1.94; p = 0.011), though not with evidence-based best practices met percentage score (p = 0.059). Outdoor play total score ranged from 43.5% to 80.2%, while evidence-based practice met percentage scores varied from 0% to 53.7%. For outdoor play, the SDOH score significantly correlated with both total percentage score (b = 3.62, 95% CI = 2.55, 4.70; p <0.0001) and evidence-based practice met percentage scores (b = 4.86, 95% CI = 2.92, 6.79; p <0.0001). Lastly, screen time total score ranged from 64.7% to 84.0%, with evidence-based practice met percentage scores ranging from 18.8% to 55.6%. SDOH score was significantly associated with total percentage score (b = 1.44, 95% CI = 0.36, 2.53; p = 0.009), but was not statistically associated with evidence-based practice met percentage scores (p = 0.060).

#### Child disability diagnosis as a moderator for associations of SDOH and family-level health practices

For the total percentage score, there were significant interactions (p < 0.10) between SDOH score and child disability diagnosis for child nutrition practices, as well as outdoor play practices. Further investigation into the interaction revealed that SDOH metrics were significantly associated with higher scores in children’s nutrition for families with TD children only (p<0.0001; Table 3). Additionally, SDOH metrics showed a significant association with higher scores in outdoor play for families with TD children (p<0.0001) and autistic children (p = 0.022), but no significant association was observed with families of young children with DS (p=0.760; Table 3). For evidence-based practice met percentage scores, there were significant interaction (p < 0.10) between SDOH score and child disability diagnosis for child nutrition, PA, and outdoor play practices. Further investigation into these interactions revealed that SDOH metrics were significantly associated to higher scores for families with TD children only for child nutrition (p<0.0001), PA (p=0.006), and outdoor play practices (p<0.0001; Table 3).

**Table 3.**
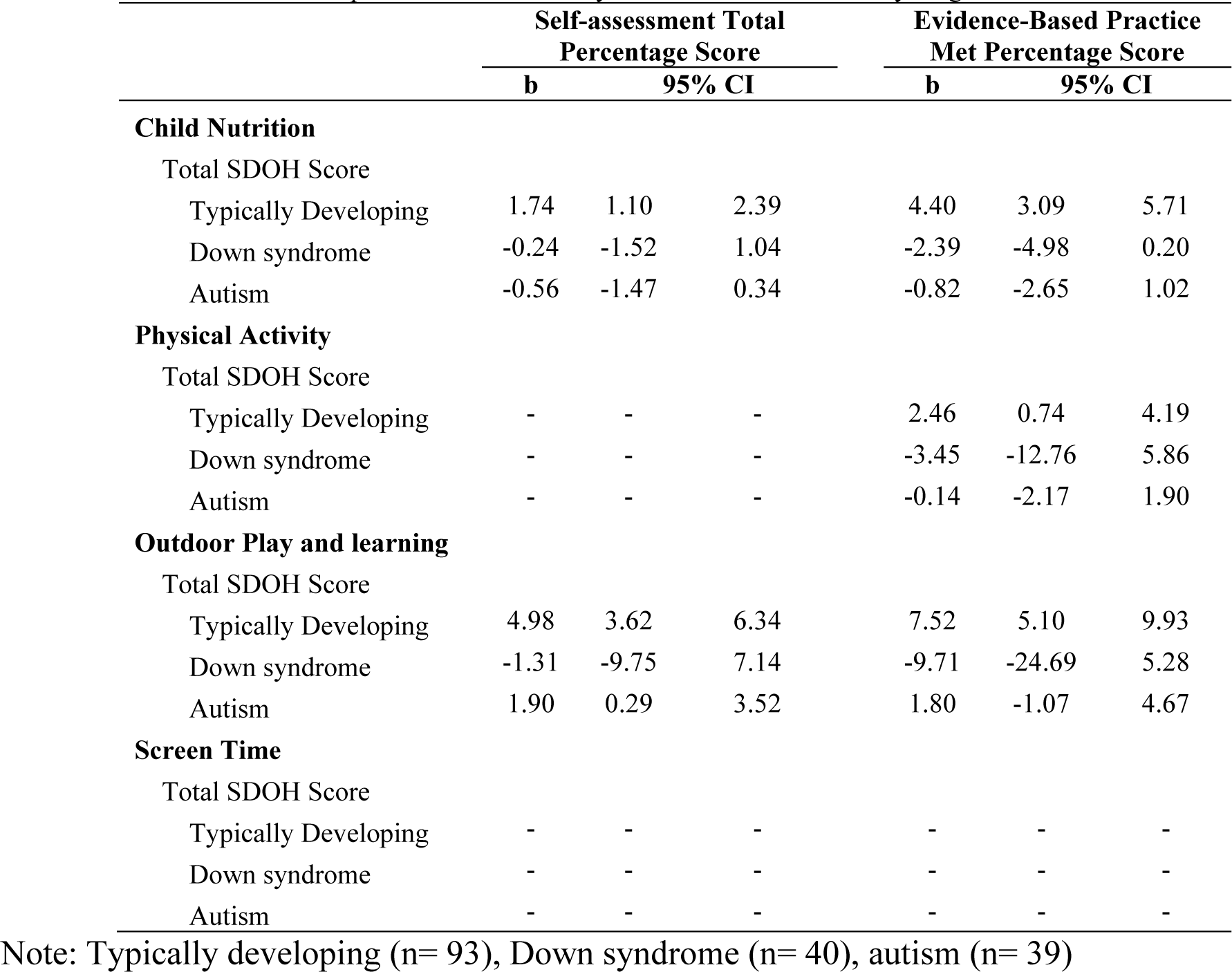
Association between child nutrition, physical activity, outdoor play and learning, screen time evidence-based practices and SDOH by child intellectual disability diagnosis.

## Discussion

Although the impact of SDOH on health outcomes and health inequalities has been well documented, no study has explored their influence on caregiver health practices in the home environment among families of children with and without IDD. The current study sought to explore the influence of SDOH on child nutrition, PA, outdoor play, and screen time among a sample of caregivers with 2-6 years old children in three groups, TD, DS, and autism, to help inform research intervention and implementation efforts in the 2-6 years old age group.

The results of this study revealed three main points. First, TD children have more advantageous SDOH factors than children with DS and autistic children. The most unfavorable SDOH factors were seen among the DS group, specifically for food insecurity, language barriers related to health information and resources, difficulty understanding resources, transportation issues, and delaying care for the child. Second, SDOH score was significantly associated with the total percentage scores (i.e., 0% is least engaging and 100% is most engaging in evidence-based practices) for all four family-level health practices (e.g., child nutrition, PA, outdoor play, and screen time), and with the percent of best practices met (i.e., 0% is not fully engaging and 100% is fully engaging in evidence-based practices) for two family-level health practices (e.g., child nutrition and outdoor play). Third, the child’s IDD diagnosis was identified as a moderator for the association between SDOH and child nutrition and outdoor play practices. In families with TD children, SDOH scores were significantly associated with child nutrition, PA, and outdoor play.

### Unfavorable SDOH factors

Families of children with IDD face many health disparities, including health care utilization (i.e., preventative services), access to care (28), higher healthcare cost (e.g., specialty services, emergency department, hospitalization) (19), poverty, food insecurity, access to educational resources, healthcare access, transportation, and social support (14). The current study corroborates these health disparities when comparing families of children with and without IDD. Our results show that caregivers of children with DS have more unfavorable SDOH factors, notably in four of the five SDOH constructs, economic stability, education access and quality, healthcare access, and residential environment. Families of autistic children have more unfavorable factors under healthcare access and social context and support. Unfortunately, caregivers of children with DS and autism reported dissatisfaction and difficulty with healthcare access.

Caregivers of children with DS were more likely to face food insecurity; they worried that the food would run out before they could buy more. Also, they experience language barrier when trying to find information or resources to support their child’s health. Although caregivers of children with DS in this study were more likely to have a high school diploma or higher education and a higher household income, which are considered favorable SDOH factors, studies have shown that children with developmental needs have higher healthcare costs, which negatively impacts the household’s income (29, 30).

While caregivers of children with DS may report a high educational level and household income, healthcare expenses associated with the child diagnosis reduces the household economic condition and makes the family more prone to income related issues such as food insecurity. The caregiver’s educational level does not explain the reported language barriers around finding health resources for the child’s wellbeing. Perhaps, these barriers are not a function of the caregiver’s level of education but are related to the healthcare provider’s knowledge (e.g., educational training), beliefs about and attitudes around children with IDD. Healthcare training deficient in IDD and low participation in research are two key areas responsible for the gap in knowledge between caregiver and healthcare provider. Many medical schools do not include disability in their training program (31); this creates a gap between what healthcare providers can offer and what families of children with IDD need. Consequently, providers do not have the necessary information or are unable to communicate it in a way that resonates with families. Thus, leaving the needs of children with IDD overlooked and unmet (32).

A significant percentage of physicians (82.4%) rate the quality of life of people with disability as worse, and only 43% “strongly” agree to welcoming patients with disability to their practice (33). Our results revealed that 37.5% of caregivers of children with DS delay care for their child because they were “nervous about seeing a healthcare provider.” People with IDDs, such as DS, are underrepresented in research (15), and the quantity and quality of health information needed to address their health needs is lacking. The increased participation in research by members of the DS community and other IDD members can help fill this existing knowledge gap. Other recommendations to address the needs of people with IDDs include improved communication strategies (e.g., health passport), a disability coordinator to improve patient care, invite persons with disabilities to participate in key conversations (e.g., ethics committees, hospital policy discussion), and increased IDD education for healthcare professionals and medical school students (34).

Children with DS have co-morbidities and need consistent and adequate access to healthcare. Although 75% of their caregivers report being satisfied with healthcare access, they are more likely to delay care for multiple reasons including “distance to healthcare provider” and “nervous about seeing a health care provider.” Moreover, lack of transportation has kept families of children with DS from key appointments (e.g., medical, work, and groceries) more frequently than families with autistic or TD children. Similarly, caregivers of autistic children report being dissatisfied with healthcare access and resources. Particularly, they are the most dissatisfied with access to healthcare services, social services programs, and health-related resources. They report not receiving adequate health information and resources for “behavior management,” “social and emotional development,” and “sleep habits.” This finding is supported by Graaf, Annis (35) who reported that parents of children with special needs who have developmental, behavioral, and emotional concerns (e.g., autistic children) need more support services compared to those who do not have those additional concerns. Additionally, caregivers’ dissatisfaction with healthcare access and services may be due to the complexity of the disorder. Autism or autism spectrum disorder covers a spectrum of symptoms and can be a complex disorder to treat. Families need coordinated care services due to the complexity of care (e.g., emotional, behavioral, medical) associated with treating autistic children (36).

### SDOH and family-level health practices

Among the four family-level health practices, there is no evidence in our sample of families fully engaging in evidence-based practices. Caregivers report that their children engaged the most in outdoor play and the least in PA. Further analysis of association revealed SDOH score is significantly associated with total percentage scores for all four family-health practices, and only for child nutrition and outdoor play for evidence-based practice met percentage score. The higher the SDOH score (i.e., more favorable factors), the higher the evidence-based practice met percentage score. These findings further support the idea that SDOH affects health quality.

As previously noted, outdoor play is associated with higher PA levels (37). In a study of children and youth with special health care needs, children did not meet the daily 60 minutes PA recommendation by the American Academy of Pediatrics and parents identified some SDOH factors as barriers to PA, including finances and being unable to pay for adaptive equipment (38). Yazdani, Yee and Chung (39) reported similar barriers including perceived lack of time for PA, lack of reliable transportation, no program that can accommodate the child’s disability, neighborhood safety, child’s behavior, and the child’s developmental delay.

### SDOH, family-level health practices, and child diagnosis

Child diagnosis was found to be a moderator for the association between SDOH and family-level health practices. SDOH score is significantly associated with higher total percentage scores in child nutrition and outdoor play for families with TD children. In contrast, there is an inverse relationship between SDOH score and percentage of evidence-based practices met score for child nutrition, PA, and outdoor play in families of children with DS. No interactions were found between total percentage score and PA and screen time, and evidence-based practices met percentage score and screen time.

#### Limitations

The purpose of this study was to explore the influence of SDOH on child nutrition, PA, outdoor play, and screen time practices among caregivers to children with and without disabilities. This study highlights an important association between the SDOH and health practices among understudied populations. However, these findings are subject to three limitations. First, the generalizability of the study results is subject to certain limitations. For instance, our study sample is not representative and lacks sufficient diversity of race, ethnicity, education, income, and relationship status among participants. Also, the sample sizes of the DS and autism group, although equal, were smaller than that of the TD group. So, the findings must be interpreted carefully. Second, our SDOH mean score was high with a relatively small standard deviation, which again highlights limited diversity in our study sample. Third, online surveys are subject to sampling bias because certain groups of people may be overrepresented or underrepresented in the data. Future research should recruit a larger and more diverse sample for race, ethnicity, education, income, relationship status, and IDD status.

## Future Research and Direction

This research’s findings have identified questions in need of further investigation, opportunities for healthcare providers, and policy implications. In this study, families of children with DS had more unfavorable SDOH factors. More research is needed to confirm these findings and understand why families of children with DS and autistic children are disproportionately affected in healthcare access and health literacy. Healthcare providers should consider the specific needs of their audience when sharing health information and clearly communicate in a way that the patient can understand and implement it. Also, a concerted effort to educate healthcare providers in all healthcare settings about IDDs and disabilities is urgently needed. This education might help address difficulties families of children with IDDs experience when accessing healthcare, improve their comfort level around healthcare personnel, and increase their health literacy. Policies at the local, state, and national level are needed to support these efforts and improve the quality of care all patients receive, especially those who are experiencing healthcare inequities with the current system. Equally, researchers should aim to recruit people with IDDs in their studies so we can increase our understanding of their health behaviors, factors that impact their health, and in turn help improve the health services they receive.

In summary, this study shows that families are struggling to meet evidence-based practices and SDOH influence primary caregivers’ health-related practices around child nutrition, PA, outdoor play, and screen time. Child’s IDD diagnosis can help explain the relationship between SDOH and child nutrition and outdoor play. Future research must prioritize SDOH in health interventions, especially in families of children with DS and autistic children to understand their varied impacts in this context.

## Practical application

SDOH influence children’s nutrition, PA, outdoor play, and screen time through the primary caregiver’s health-related practices. Researchers, educators, and healthcare providers should consider SDOH factors that families may face when working to improve their child’s health behavior.

## Data Availability

The datasets generated and/or analyzed during the current study are available from the corresponding author on reasonable request.

## Acknowledgments

“We express our sincere gratitude to the families who took part in this study and thank them for their commitment and time. Thank you to the University of North Carolina Chapel Hill’s Intellectual and Developmental Disabilities Research Center Registry Core funded by the National Institute of Child Health and Human Development (P50HD103573) for assistance in disseminating information about the project to families. Thanks also to Vishwa Patel, Jamie Halula, and Derek Hales for their assistance editing and testing the online survey. In memoriam to Dr. Dianne Stanton Ward, whose profound influence and support has left an enduring mark on the development and progression of our research.

## CRediT Authorship

Phoebe P. Tchoua: Writing – review & editing, Writing – original draft, Methodology, Formal analysis, Conceptualization.

Emily C. Clarke: Writing – review & editing,

Heather Wasser: Writing – review & editing

Seema Agrawal: Writing – review & editing,

Rebecca Scothorn: Writing – review & editing

Kelsey Thompson: Writing – review & editing

Michaela A. Schenkelberg: Writing – review & editing,

Erik A. Willis: Writing – review & editing, Writing – original draft, Supervision, Methodology, Investigation, Data curation, Conceptualization.

